# Procoagulant effect of phosphatidylserine exposed to extracellular vesicles, blood cells and endothelial cells in patients with aortic stenosis

**DOI:** 10.1101/2024.06.11.24308758

**Authors:** Zhaona Du, Haiyang Wang, Yibing Shao, Wei Wu, Dongxia Tong, Fangyu Xie, Jihe Li, Wei Xia, Yujie Zhou

## Abstract

**Background:** The mechanism of thrombotic complications in patients with aortic stenosis (AS) is unknown. Our aim was to evaluate the levels of phosphodiesterase (PS) in blood cells, endothelial cells (ECs), and extracellular vesicles (EVs) and its procoagulant activity (PCA) in different degrees of AS.

**Methods:** Exposed PS in blood cells, ECs and EVs were analyzed by flow cytometry. PCA was evaluated by clotting time (CT), intrinsic factor Xa (FXa), extrinsic FXa, thrombin and fibrin formation assays. We also evaluated the inhibitory effects of lactadherin (Lact) and anti-tissue factor (anti-TF) on PCA in severe AS patients.

**Results:** Our results demonstrated that positive phosphatedylserin (PS^+^) with total EVs, platelet EVs (PEVs), positive tissue factor EVs (TF^+^EVs), and endothelial-derived EVs (EEVs) levels were significantly higher in mild to severe AS than controls. Patients with AS had significantly higher percentages of PS^+^ red blood cells (RBCs), white blood cells (WBCs), platelets (PLTs) and ECs compared to controls. In addition, we further confirmed that PS^+^ blood cells, ECs and EVs significantly contributed to shortened CT and dramatically increased FXa, thrombin and final fibrin generation in mild to severe AS compared to controls. Furthermore, in severe AS, lactadherin significantly inhibited PCA of PS exposure in blood cells, ECs and EVs, whereas anti-TF had no effect.

**Conclusion:** Our study revealed a previously unrecognized association between exposed PS levels on blood cells, ECs and EVs and PCA in AS. Lactadherin promises to be a new therapy by blocking PS to prevent thrombosis in AS patients.

## Introduction

As the age grows, the incidence of Aortic stenosis (AS) is increasing. Study found that AS occurs in 10% of people over the age of 80 and is often associated with a poor prognosis^1^. Thromboembolic complications are major determinants of prognosis and quality of life in AS patients^1, 2^. AS-associated thromboembolic complications can lead to thromboembolic stroke, myocardial infarction, pulmonary embolism and heart failure^1, 3^. Thromboembolic complications in patients with severe AS is a major risk factor for stroke and death^1, 4^. The pathophysiology of thromboembolism in AS is multifactorial. Hypercoagulable state of blood is the potential pathogenesis^3^. Previous studies showed that there was no significant difference between anticoagulation and antiplatelet therapy in terms of cardiovascular thromboembolic events^5, 6^. The exact mechanism of the formation of hypercoagulable state in AS has not been clarified, and the optimal anticoagulation mechanism has not been perfected, so further studies on the mechanism of the prethrombotic state in AS patients are needed.

It is well known that inflammation, infection, tumors and immunodeficiency can activate circulating blood cells and inflammatory cells^7-9^. Activated blood cells such as erythrocytes, leukocytes and platelets have been shown to induce a prethrombotic state in the body^10-12^. This phenomenon has been observed in diseases such as coronary atherosclerosis^13^ and atrial fibrillation^14^. There is increasing evidence that erythrocytes, leukocytes and platelets are activated in AS^1, 15, 16^. In addition, other evidence showed that ECs were activated in AS^17^. Previous studies found that high shear in AS promoted the activation of circulating cells and ECs^18^. However, the extent of activation of blood cells and ECs in these patients is not fully understood.

Circulating EVs are plasma membrane-derived vesicles shed by various types of activated or apoptotic cells, including PLT, monocytes, ECs, erythrocytes and granulocytes^19^. Recent evidence suggest that they may play important functions in cell-cell interactions, homeostasis and pathogenesis of a number of diseases^19^. In addition, EVs play a role in the procoagulant activity in diseases by expressing PS, tissue factor (TF) and other coagulation factors^20^. PS and TF are known to be associated with the prethrombotic state in cardiovascular diseases^21, 22^. However, there are few studies on EVs levels in patients with AS. It is not entirely clear the level changes of PS and TF in AS and their role in PCA.

In this study, we investigated the differences in the levels of EVs released from blood cells and ECs between mild to severe AS and controls. We also analyzed the levels of PS exposure in blood cells, ECs, and EVs in AS. In addition, we evaluated the contribution of PS exposure to PCA and further analyzed the disturbance of lactadherin and anti-TF on coagulation activity in AS patients. Our study may contribute to the discovery of new therapeutic targets to effectively intervene in the thrombotic complications of AS and reduce the risk of death.

## Methods

### Study population

In our study, patients diagnosed with AS by thoracic echocardiography were continuously selected for the study from the Heart Center of (Qingdao Hospital) University of Health and Rehabilitation Sciences from November 2020 to September 2022. A total of 166 patients met the inclusion and exclusion criteria, including 55 mild AS, 51 moderate AS and 60 severe AS patients. Fifty healthy volunteers were recruited as healthy controls in the same time.

Exclusion criteria for all patients were congenital heart disease, bicuspid aortic valve stenosis, history of aortic valve surgery, history of stroke, history of heart failure, history of thromboembolism, acute and chronic inflammatory diseases, history of surgical procedure within 3 months of presentation, hepatic or renal disease, autoimmune or malignant diseases, and receiving or about to receive anticoagulant or antiplatelet therapy within 2 weeks. We graded the aortic valve stenosis according to the valve orifice area, mild stenosis was that the valve orifice area was reduced but greater than or equal to 1.5 square centimeters; moderate stenosis was that the valve area was between 1.0 and 1.5 square centimeters and severe stenosis was that the valve area was less than or equal to 1.0 square centimeters^23^.

In accordance with the Declaration of Helsinki, approval was obtained from the Research Ethics Committee of our institution, and written informed consent was obtained from all participants.

### Experimental procedures

#### Materials

We used a polyclonal antibody to human TF from American Diagnostica Inc. (Stamford, CT, USA) and labeled all monoclonal antibodies with Alexa Fluor 647 or 488. CD142 (clone HTF-1), CD41a (clone HIP8), purified CD31 (clone L133.1) and TruCount Tube from Becton Dickinson (San Jose, CA, USA) were also used. Human FXa, thrombin, and prothrombin were obtained from Haematologic Technologies. Alexa Fluor 647 or 488 conjugated lactadherin and Tyrode’s buffer containing 1 mM HEPES were prepared and filtered through a 0.22 mm syringe filter from EMD Millipore. Chromogenic substrates S-2765 and S-2238 were purchased from DiaPharma Group. Calibrated polystyrene latex beads (1.0 μm) were purchased from Sigma (UK).

#### Protein purification and marking

The ratio of lactadherin to fluorescein in this study was 1/ 1.1-1.2, and we used Alexa Fluor 647 or Alexa Fluor 488 to label lactadherin purified from milk^24^.

#### Endothelial Cell Culture and Recombinant experiments

Human umbilical vein endothelial cells (HUVECs) were cultured using ECs medium (Science Cell, San Diego, CA, USA) at 37°C in a 5% CO_2_ humidified atmosphere. ECs were processed from media containing 20% pool serum from mild AS, moderate AS, severe AS patients and healthy controls at room temperature for 24 hours. Using the flow cytometer to define PS exposure^25^.

#### Preparation and flow cytometric analysis of EVs

Blood samples were collected from controls and AS with different degrees of stenosis. Within 30 minutes after blood collection, the samples were centrifuged for 20 minutes at 1500 g. The upper plasma layer was aspirated and centrifuged again for 30 minutes at 20,000 g. We then removed the plasma supernatant, collected the remaining EV at the bottom of the test tube and stored at -80°C. Lactadherin-positive (lac^+^), lac^+^CD41a^+^, lac^+^CD31^+^41a^-^, and lac^+^CD142^+^ were used to characterize PS^+^EVs, PEVs, EEVs, and TF^+^EVs. The number of EVs of each type per microliter was calculated using a Trucount tube after accumulation of 10,000 gated events using the following formula: n = (C × Beads added) / (Beads counted × Sample volume). In the formula, ‘C’ stands for the number of positive events after subtraction of the background signal.

#### Coagulation time and inhibition assays of EVs

KC4A coagulometer was used to evaluate the PCA of EVs. We mixed EV-enriched suspension and Tyrode’s buffer in a ratio of 1:9 to obtain EV-containing suspension, and incubated 100 microliters of EV-containing suspension with 100 μl of EV-depleted plasma at 37°C for 3 min. 100 μl of warmed CaCl^2^ (25 mM) was added to start the reaction and record the coagulation time (CT). In the inhibition assay, we incubated 100 μl of EVs suspension with 50 μl of lactadherin or anti-TF for 10 min at 37 °C. The coagulation time was recorded after the addition of 100 μl of EV-free human plasma and 50 μl of warmed CaCl_2_ (50 mM).

#### Intrinsic, extrinsic FXa and thrombin formation and inhibition assays of EVs

We first performed FXase and prothrombinase activity assays on all samples. To prepare intrinsic Xa, we incubated 10 μL EVs suspension with FX (130 nm), Fixa (1 nm), thrombin (0.2 nm), and CaCl_2_ (5 mm) in FXa buffer (10 ml 1 × incubate in TBS containing 0.2% BSA) at 25° C for 5 minutes, then used EDTA (7 mm final concentration) to stop the reaction. FXa production was measured using a universal microplate spectrophotometer (PowerWave XS; Bio-Tek, Winooski, VT, USA) at 405 nm after incubation with 10 μl of S-2765 (0.8 mM). Except for EVs cultured with FX (130 nM), FVIIa (1 nM) and CaCl_2_ (5 nM), the formation of extrinsic FXa was similar to that of intrinsic FXa. In the prothrombinase assay, EVs with 0.05 nM FXa (0.05 nM), FVa (1 nM), prothrombin (1 μM), and CaCl_2_ (5 nM) were incubated in prothrombinase buffer (10 ml 1 × TBS with 0.05% BSA) for 5 min at 25°C and then stopped with EDTA. For inhibition assays, EVs were preincubated with lactadherin (128 nM) or anti-TF at their peak time at 25°C in Tyrode’s buffer for 10 minutes and then incubated with the specified coagulation factors. The formation of FXa or thrombin was assessed as described above.

#### Fibrin formation assays

Fibrin formation was evaluated by turbidity. EVs were added to recalcified (10 mM, final) EV-depleted plasma (88% EV-depleted plasma, final) in the absence or presence of lactadherin/anti-TF. A SpectraMax 340PC plate reader was used to measure fibrin production by turbidity at 405 nm.

### Statistical analysis

SPSS v27.0 software and GraphPad Prism 9.0 were used to analyze statistics. Data conforming to a normal distribution are expressed as mean ± standard deviation and were statistically analyzed using Student’s t-test or ANOVA, as appropriate. Categorical variables were compared using the χ2 test. Pearson or Spearman’s rank correlation analysis was used to express correlations between two continuous variables. P < 0.05 was considered statistically significant.

## Results

### Participants characteristics

We analyzed the clinical characteristics of 50 healthy controls and all AS patients (**Table 1**). There were no significant differences in gender, current smoking, current alcohol, hypertension, diabetes mellitus, atrial fibrillation and stroke between control and mild to severe AS patients. Age, body mass index (BMI), aortic valve area (AVA), fasting glucose, aspartate aminotransferase (AST), aspartate aminotransferase (AST), alanine aminotransferase (ALT), creatinine (Cr), triglyceride (TG), serum total cholesterol (TC) and low-density lipoprotein cholesterol (LDL-C) levels were significantly higher in AS than control. While high-density lipoprotein cholesterol (HDL-C) levels were significantly lower in AS than controls. In addition, the levels of white blood cells (WBC), neutrophils, red blood cells (RBC), platelets (PLT), fibrinogen, D-dimer (D-D) and thrombin-antithrombin complex (TAT) levels were markly higher in AS compared to controls. Conversely, prothrombin time (PT) and activated partial thromboplastin time (APTT) levels were significantly lower in all AS patients than controls. Further analysis showed that severe and moderate AS patients had significantly different levels of WBC, neutrophils, RBC, PLT, fibrinogen, D-D, TAT, PT, APTT, AST, ALT, Cr, TG, TC and LDL-c than mild AS groups.

### The percent of lactadherin-binding blood cells and ECs in mild to severe AS and controls

We analyzed the percent of lactadherin-binding blood cells and ECs by flow cytometry (**Fig. 1**). The percentage of RBC, PLT, WBC and serum-cultured ECs were significantly higher in AS patients compared to control groups. Among all AS groups, severe AS had the highest PS exposure rate of RBC, PLT, WBC and ECs.

The relationships between the percentages of total PS^+^EVs, LAC^+^ RBC, LAC^+^ PLT, LAC^+^ WBC and LAC^+^ ECs and hypercoagulable markers (D-dimer or TAT) were also analyzed (**Table 2**). In mild, moderate and severe AS patients, D-dimer (D-D) and TAT had a significant positive correlation with the percentages of total PS^+^EVs, LAC^+^PLT, LAC^+^RBC, LAC^+^WBC and LAC^+^ECs. It can be concluded that, to some extent, changes in PS exposure on circulating blood cells, ECs and EVs could lead to PCA in AS patients.

### Dynamics of PS^+^EVs/EEVs/PEVs/TF^+^EVs in mild to severe AS and controls

The total number of EVs and their phenotypic features were evaluated by flow cytometry in Figure 1. Circulating total EVs levels were significantly elevated in mild to severe AS groups compared with controls, and there was a statistically remarkable difference among AS groups (**Fig. 1**). As the degree of AS stenosis increased, EVs levels were significantly elevated, with more pronounced elevations in patients with severe AS (**Fig. 1**).

### PCA of PS^+^ blood cells, EVs and ECs in AS patients

As we predicted, from mild to severe AS, the PS^+^ EVs, RBC, PLT and WBC showed significantly shorter CT than those from controls, and the ECs pretreated with each AS group serum also had a shorter CT than the control (**Fig. 3A**). We further analyzed the coagulation activity of PS^+^ EVs, blood cells and ECs by intrinsic FXa, extrinsic FXa and thrombin generation assays (**Fig. 3 B, C**). We found that in all AS, the production of the intrinsic FXa, extrinsic FXa and thrombin production were increased in PS^+^ EVs, PLT, WBC and ECs, and the production of intrinsic FXa and thrombin were increased in RBC than in controls (**Fig. 3B, C**). To further verify whether this increase in PCA was due to PS exposure, in an inhibition assay of the severe AS group, we used lactadherin (128 nM) and anti-TF to block available PS (**Fig. 3 D, E, F, G**). We found that preincubation with lactadherin significantly prolonged the CT of PS^+^ EVs, blood cells and ECs in severe AS compared with anti-TF in severe AS and controls (**Fig. 3D**). In addition, lactadherin significantly reduced intrinsic FXa, extrinsic FXa and thrombin production in the severe AS patients, whereas anti-TF had no difference **(Fig. 3E, F, G**).

### Fibrin formation assays of EVs, blood cells and ECs

To evaluate the role of PS on fibrin production, we measured the final fibrin production of EVs, RBC, PLT, WBC and ECs in each AS group and examined the effects of lactadherin and anti-TF on the final fibrin content of severe AS group (**Fig. 4A, B**). The final fibrin production content was significantly increased in each AS group of EVs, hemocytes and ECs compared with the normal control group, and the increase was most marked in the severe AS group (**Fig. 4A**). The addition of lactadherin significantly reduced the final fibrin level in severe AS patients, whereas anti-TF had no effect on the final fibrin formation (**Fig. 4B**).

## Discussion

The levels of PS exposure in hemocytes, ECs or EVs and its role in AS coagulation is unclear. In this study, our data showed that the percentages of PS^+^ blood cells and ECs were significantly higher in mild, moderate and severe AS groups. D-D and TAT were significantly positively correlated with PS levels and increased with increasing PS levels. In addition, the AS group had a significantly shorter CT and a significant increase in FXa, thrombin and final fibrin formation as the level of PS exposure increased. The procoagulant inhibition test showed that lactadherin significantly prolonged CT, decreased the levels of procoagulant enzyme complexes and final fibrin formation. This suggests that the levels of PS exposure in EVs, RBCs, PLT, WBCs and ECs play a non-negligible role in prothrombinase assembly and fibrin formation in AS patients.

Study found that high shear stress was an inducer of blood cell activation in the development of arterial disease^26^. Diehl et al^27^ demonstrated a correlation between shear stress and platelets or leukocytes in AS patients. In addition, some studies found that the presence of high shear stress in AS activated the expression of ECs^17, 28^. In our study, the activation numbers of RBC, PLT, WBC and ECs were significantly higher in all AS groups than controls, which is consistent with previous findings. This was mainly caused by altered hemodynamics due to stenotic aortic orifices and thickened valves, resulting in ECs and circulating blood cells activated by abnormal shear forces^12, 17, 29^. It is noteworthy that our study found that among all AS patients, the number of activated RBCs, PLT, WBCs and ECs was significantly higher in severe AS patients than in mild and moderate AS patients. In addition, our study further demonstrated that EVs and PS exposure of these activated cells and ECs were significantly elevated in AS. From this, we can conclude that the high shear stress generated in AS patients affected the activation and release of hemocytes and ECs, and further increased EVs and PS levels, the degree of valve stenosis played a crucial role in the activation of blood cells and ECs.

EVs are small cell membrane vesicles of 0.1-1 μm released by apoptosis, abnormal activation of blood cells and endothelial cells of membrane remodeling, during states of inflammation, hypoxia and high shear levels, EVs shedding can increase^22, 30^. Research has shown that high shear stress in AS is associated with the release of circulating leukocyte, platelet and endothelial cell derived EVs^27, 31^. In this study, we demonstrated that the total levels of PS^+^ EVs, EEVs, PEVs and TF^+^EVs were significantly higher in all AS groups than in healthy controls. Previous studies have shown that the levels of EVs from WBC, PLT and ECs were increased in severe AS compared to controls, which was consistent with our findings^27^. In addition, studies have shown that high levels of EVs were a major contributor to hypercoagulability^25^. Which is consistent with our findings. This is mainly because circulating EVs provide an additional procoagulant phospholipid surface for activation of the coagulation cascade reaction^32^. Therefore, it can be concluded that the shear stress caused by AS may contribute to the coagulation process by activating the generation of EVs from hemocytes and ECs. More importantly, the PCA of EVs in AS is highly dependent on the level of PS exposure^32^.

On the plasma membrane, PS was preferentially found in the inner leaflet^33^. During biological processes such as apoptosis and activation, PS could be exposed on the cell surface^34^. Research has shown that the increased levels of PS^+^ ECs occur in some diseases^35^. In AS, leukocytes, lymphocytes, neutrophils, macrophages and monocytes were abnormal activation, PS exposure on the plasma membrane surface was increased^36, 37^. Our study showed that patients with mild AS to severe AS showed higher levels of PS^+^ RBC, WBC, PLT and ECs were significantly higher in AS groups than controls. This was similar to previous findings. In addition, studies have confirmed that PS exposed to blood cells, ECs and EVs can provide sites for the coagulation cascade reaction, which promoted the process of coagulation^35-37^. This further confirms our findings. Thus, we can easily conclude that activated hemocytes, ECs and EVs-exposed PS played a key role in PCA in AS patients^32^.

Previous studies have shown that thrombin generation was increased in AS^38^. In our study, we identified the role of PS exposure in thrombin generation in AS patients and additionally analyzed the relationship between PS exposure and markers of high cohesion. Early studies have shown that AS patients had increased levels of coagulation markers^1^, which is consistent with the results of our study. However, our study further explained the impact indicators of elevated levels of hypercoagulable markers in AS. Our findings suggested that PS played a key role in influencing the levels of coagulation markers and the final fibrin formation in AS patients. The rate of PS exposure of total EVs, blood cells and ECs were positively correlated with D-D and TAT in AS patients, which further confirmed the promoting role of PS exposure in AS thrombosis.

In previous studies, PS has been shown to promote a hypercoagulable state in the body under pathological conditions^1^. Our study further demonstrates the role of PS in AS, compared with the control group, the CT of PS^+^ EVs, RBC, PLT, WBC, and ECs in AS patients was shortened, the content of plasminogen complex was increased, and the body was in a hypercoagulable state. In addition, TF may also played a role in the procoagulant process^1^. However, the contribution of PS and TF to the procoagulant process in AS is unclear. For further analysis, we used anti-TF antibody and lactadherin to disrupt the exposure levels of TF and PS. Our results showed that the inhibitory effect of lactadherin on PS led to a significant decrease in the coagulation activity of PS in the samples, whereas the coagulation activity was not significantly altered in the group with the inclusion of the anti-TF antibody. This did not fully explain our finding of increased TF^+^ EVs in AS, but our study also found that TF^+^ EVs were a smaller proportion of total EVs. Previous studies also did not find a critical role for TF^+^ EVs in PCA^39, 40^. This could be caused by the fact that cycling TF are mostly in an encoded state and often cycle in a coagulated inactive or cryptic form, clusters of PS can be used to decode for TFs and increase TF activity^32, 41^. Another finding that TF was in contact with the circulation but did not induce significant coagulation suggested that TF circulated in a coagulation-inactive or cryptic form in most cases and rarely played a role, which further supported our inhibition experimental results^41, 42^. Thus, PS played a major role in the high procoagulant activity in AS patients.

## Data Availability

All data produced in the present study are available upon reasonable request to the authors

## 4. Conclusions

In conclusion, our results showed that AS patients had a hypercoagulable state compared to controls. Elevated levels of PS exposure in blood cells, ECs and EVs may lead to hypercoagulability in AS patients. PS levels and coagulation activity were significantly higher in AS patients as the severity of AS increased. In addition, the addition of lactadherin to block PS resulted in a significant decrease in PCA in AS patients, thus lactadherin is expected to be a new therapy for thromboprophylaxis in AS patients.

## 5. Limitation

One of the main limitations of our study is that it is cross-sectional, a single-center study. In addition, our sample size was small and externally there is a need for further studies with multi-center, large samples.

## Author contributions

Z.D., H.W., Y.S., J.L., W.X. and Y.Z. designed the experiments, analyzed data, and wrote the paper. W.W. and D.T. performed the experiments. F.X. helped with the experiments.

## Acknowledgments

We would like to specially thank Qingdao Municipal Hospital Heart Center and Qingdao Municipal Hospital Central Laboratory, Qingdao, China.

